# Evaluation of a Novel Therapeutic Repetitive Transcranial Magnetic Stimulation Technique Optimized for Increased Accessibility in Major Depression

**DOI:** 10.1101/2020.06.15.20132092

**Authors:** Jean-Philippe Miron, Helena Voetterl, Linsay Fox, Molly Hyde, Farrokh Mansouri, Sinjin Dees, Ryan Zhou, Jack Sheen, Véronique Desbeaumes Jodoin, Arsalan Mir-Moghtadaei, Daniel M. Blumberger, Zafiris J. Daskalakis, Fidel Vila-Rodriguez, Jonathan Downar

**Author notes:** Corresponding author, 900 rue Saint-Denis, Office R09-422, Montréal, Québec, Canada, H2X 0A9.

## Abstract

Although effective in major depressive disorder (MDD), repetitive transcranial magnetic stimulation (rTMS) is costly and complex, limiting accessibility. We thus tested the feasibility of a novel rTMS protocol optimized for scalability. Several novel techniques were explored, such as an open-room setting, large non-focal parabolic coils, and cost-saving custom-built coil arms. We employed a low-frequency (LF) 1 Hz stimulation protocol (360 pulses per session), delivered on the most affordable FDA-approved device. MDD participants received an initial accelerated rTMS course (arTMS) of 6 sessions/day over 5 days (30 total), followed by a tapering course of daily sessions (up to 25) to decrease the odds of relapse. The self-reported Beck Depression Inventory II (BDI-II) was used to measure severity of depression. Forty-eight patients completed the arTMS course. No serious adverse events occurred, and all patients reported manageable pain levels. Response and remission rates were 35.4% and 27.1% on the BDI-II, respectively, at the end of the tapering course. Repeated measures ANOVA showed significant changes of BDI-II scores over time. If rTMS could be delivered for lower cost at higher volume, while preserving efficacy, safety and tolerability, it could warrant further investigation of this treatment as a first-line intervention in MDD.

## 1. INTRODUCTION

Major depressive disorder (MDD) is a common and disabling illness. Up to 50% of patients experience a chronic or recurrent course, and 30 to 40% develop treatment-resistant depression (Lam et al., 2016). Furthermore, although antidepressants offer high convenience and simplicity of administration, discontinuation rates approach 50% after 3 months of use due to concern over side effects and non-response (Kennedy et al., 2016). Alternative treatments are therefore needed.

Repetitive transcranial magnetic stimulation (rTMS) is recognized as an effective intervention in MDD, with recent studies and meta-analyses reporting response and remission rates of up to 50-55% and 30-35%, respectively (Milev et al., 2016). rTMS has an advantageous side effect profile, with lower discontinuation rates than medication (∼5% vs ∼25%) (Milev et al., 2016). Unfortunately, its widespread adoption is impeded by several obstacles that limit clinical accessibility.

The main issue concerns high acquisition and operation costs, with average cost per remission estimated to be of up to $6,146 in the US (Mendlowitz et al., 2019). This is partly a result of the equipment needed to deliver high-frequency (HF) protocols, as well as the need for continuous 1:1 technician-patient supervision during treatment. Technical challenges stem from the complexity of the treatment, such as the need for precise positioning of the widely used figure-of-eight (Fo8) focal coils over the target region.

Another area that remains to be refined is treatment course optimization. The current treatment paradigm forces patients to travel to a treatment center every day for 6 weeks in order to receive a full 30 session course. This can be discouraging, especially for working individuals and those who have families. Several studies have explored “accelerated” rTMS (arTMS), whereby multiple stimulation sessions are delivered per day to shorten the overall treatment course duration. Several large trials of arTMS have consistently reported similar or better remission and response rates than conventional once-daily rTMS (Baeken, 2018; Baeken et al., 2013; Dardenne et al., 2018; Desmyter et al., 2016; Duprat et al., 2016; Fitzgerald et al., 2018; George et al., 2014; Holtzheimer et al., 2010; McGirr et al., 2015; Schulze et al., 2017; Sonmez et al., 2019; Williams et al., 2018). Furthermore, the total cumulative number of sessions needed to achieve maximal effect is still debated. So far, findings from large rTMS trials have suggested a plateau in clinical response at ∼30 sessions, on average (Perera et al., 2016). This has recently been challenged by a secondary analysis (Kaster et al., 2019) from the THREE-D trial (Blumberger et al., 2018), demonstrating differences in patients’ response trajectories to rTMS. Indeed, some participants were still showing signs of improvement at 30 sessions (not reaching a plateau), suggesting that a treatment extension might be needed in some individuals.

To address these issues, we developed a novel rTMS technique optimized for maximum practicality, scalability and effectiveness, while minimizing costs. We employed a safe, well-tolerated low-frequency (LF) 1 Hz stimulation protocol on the lowest cost FDA-approved devices, delivered via large parabolic coils held by low-cost custom arms enabling simple yet accurate placement, in an open-room setting enabling supervision of multiple simultaneous sessions. We also tested the effects of 1 Hz arTMS followed by a tapering course of once-daily treatment. These technical refinements are designed to enable the provider to accelerate, increase and maintain treatment response beyond what ‘standard of care’ rTMS can achieve, while achieving higher patient volumes at lower cost.

We hypothesized that our novel rTMS technique would be safe, well-tolerated and effective, while allowing cost-saving opportunities and therefore increasing accessibility.

## 2. METHODS

### 2.1 Participants

We conducted a prospective, single-arm, open-label feasibility study. Participants were recruited after referral to the Poul Hansen Family Centre for Depression neurostimulation specialty clinic, located at the Toronto Western Hospital, an academic healthcare centre which is part of the University Health Network (UHN) in Toronto, Canada. Adult (18-85 years of age) outpatients were included for study participation if they 1) had a Mini International Neuropsychiatric Interview (MINI) confirmed MDD diagnosis (single or recurrent episode) and 2) maintained a stable medication regimen from 4 weeks before treatment start to the end of the study. Exclusion criteria were: 1) history of substance dependence or abuse within the last 3 months; 2) concomitant major unstable medical illness; 3) cardiac pacemaker or implanted medication pump; 4) active suicidal intent; 5) diagnosis of any personality disorder as assessed by a study investigator to be primary and causing greater impairment than MDD; 6) diagnosis of any psychotic disorder; 7) any significant neurological disorder or insult (including, but not limited to: any condition likely to be associated with increased intracranial pressure, space occupying brain lesion, any history of seizure confirmed diagnostically by neurological assessment [except those therapeutically induced by ECT], cerebral aneurysm, Parkinson’s disease, Huntington’s chorea, dementia, stroke, neurologically confirmed diagnosis of traumatic brain injury, or multiple sclerosis); 8) if participating in psychotherapy must have been in stable treatment for at least 3 months prior to entry into the study (with no anticipation of change in the frequency of therapeutic sessions, or the therapeutic focus over the duration of the study); 9) any clinically significant laboratory abnormality in the opinion of the investigator; 10) a dose of more than lorazepam 2 mg daily (or equivalent) currently (or in the last 4 weeks) or any dose of an anticonvulsant due to the potential to limit rTMS efficacy and 11) any non-correctable clinically significant sensory impairment. All participants provided informed consent and this study was approved by the Research Ethics Board of the University Health Network.

### 2.2 Study design and procedure

Treatment was delivered in an open room setting, allowing up to 4 participants to receive treatment simultaneously, with the help of one or two technicians (**Figure 1**). This was facilitated by an easy-to-use coil placement system, with the Anchored Articulating Arm (AAA) concept at its core (**Figure 2**). AAAs are made out of readily available camera tripod equipment and are anchored at their base to the corner of the treatment chair. Coils are held securely by a clamp locking it to the AAA, maintaining position until treatment has been completed, at which time they can be easily repositioned out of the way.

**Figure 1:**
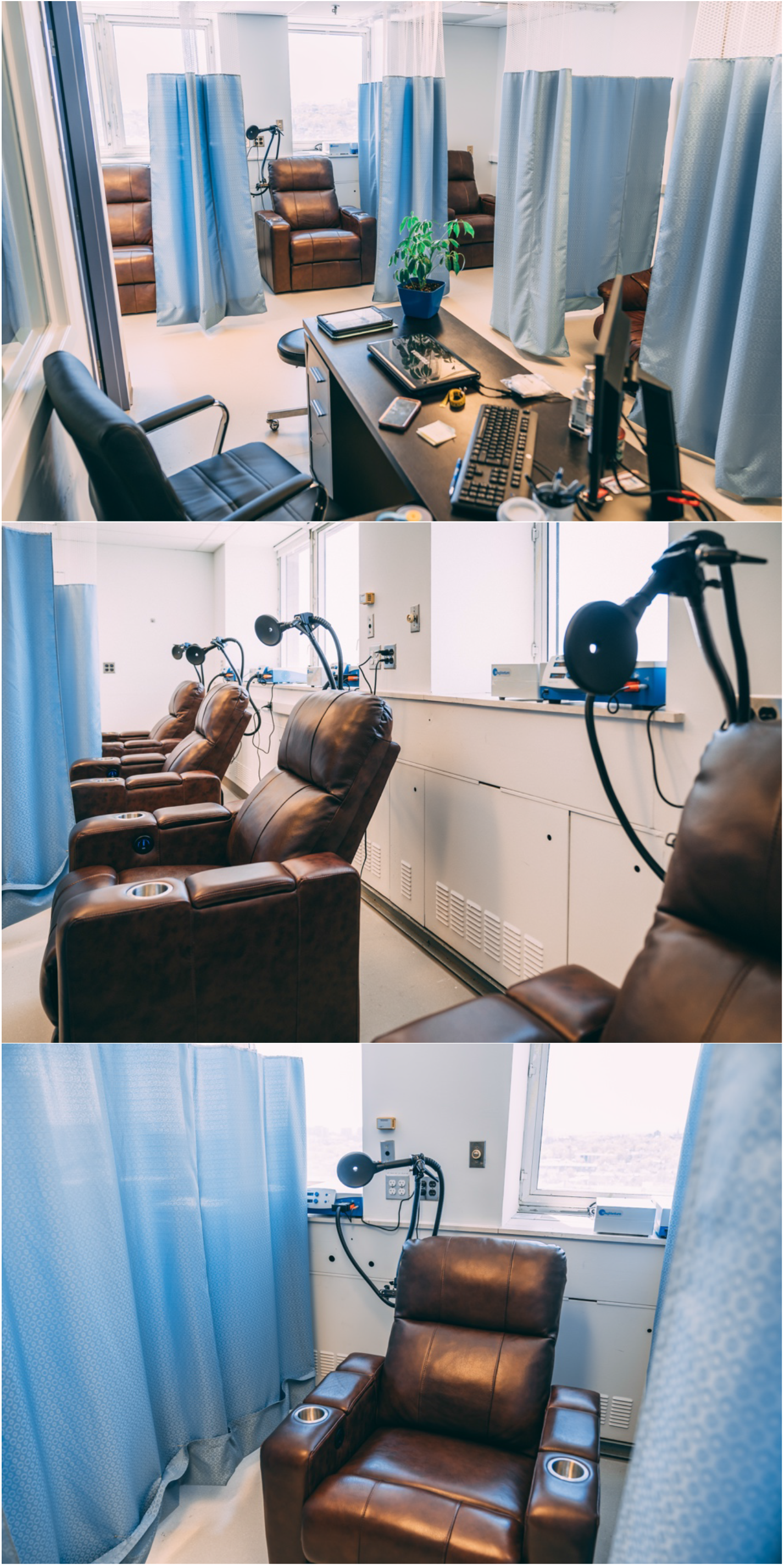
Clinical setup - 4 recliners equipped with AAAs holding rTMS coils, connected to the rTMS stimulators. Treatment was delivered in an open room setting, allowing up to 4 participants to receive treatment simultaneously, with the help of one or two technicians. AAA = Anchored Articulating Arm

**Figure 2:**
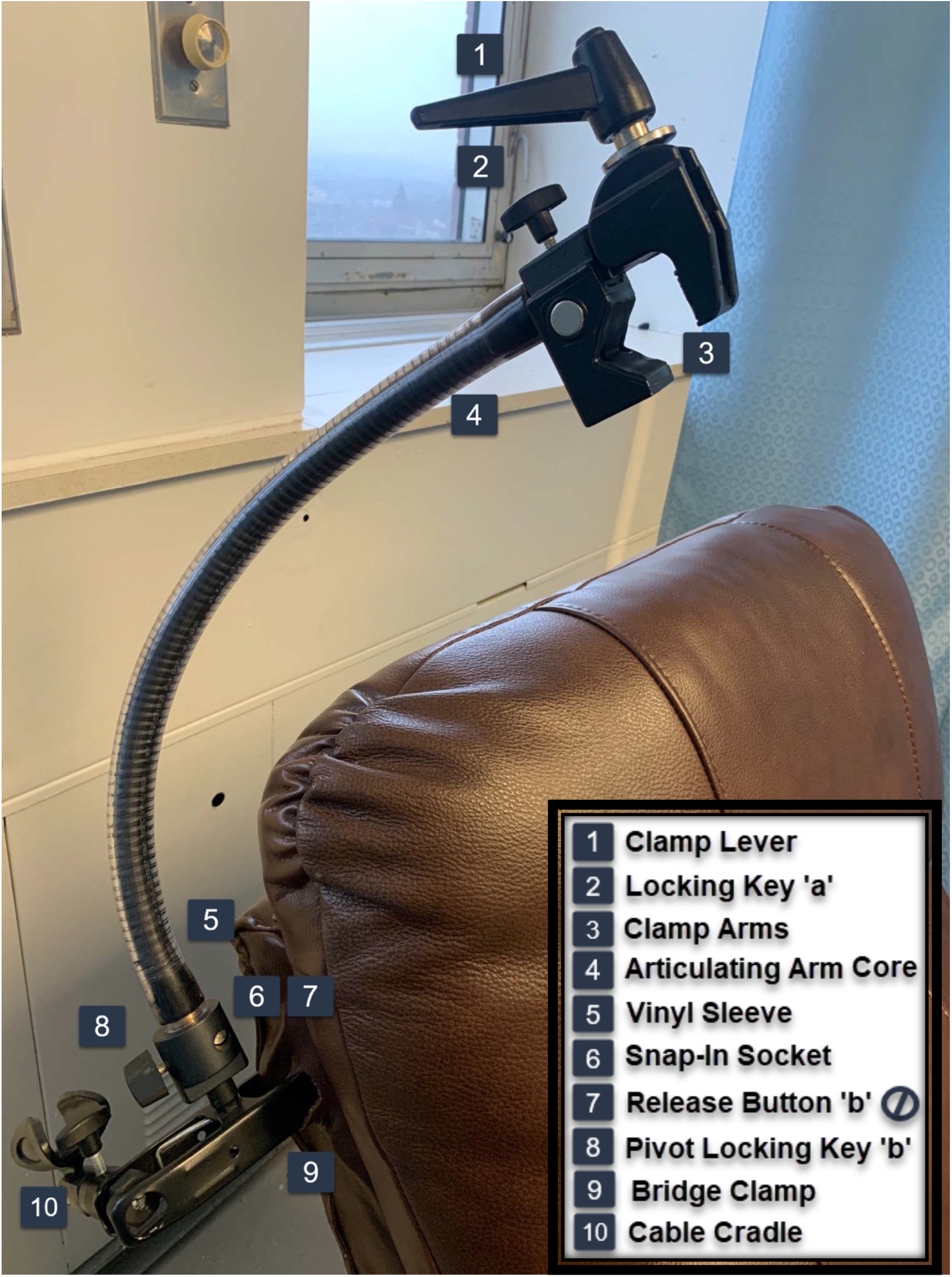
Anchored Articulating Arm (AAA) schematic - AAAs are made out of inexpensive camera tripod equipment and are anchored at their base to the corner of the treatment chair. Coils are held securely by a clamp locking it to the AAA, maintaining position until treatment has been completed, at which time they can be quickly twisted out of the way.

rTMS was delivered through MagPro R20 stimulators equipped with parabolic MMC-140 coils (MagVenture, Farum, Denmark). We recently published a case series on the safety, tolerability, and effectiveness of those coils in MDD (Miron et al., 2019). The resting motor threshold (rMT) was determined according to standard clinical practice, with the additional need to use the coil’s middle ring for stimulation, where the electromagnetic field strength is at its highest (McClintock et al., 2017; Miron et al., 2019). We used a previously published 1 Hz rTMS protocol (60 s on and 30 s off, 6 trains, 8.5 min total stimulation time, 360 pulses/sessions, 120% rMT) over the right dorsolateral prefrontal cortex (DLPFC) with the coil centred on the F4 EEG location (right-flipped adjusted BeamF3) (Brunelin et al., 2014; Miron et al., 2019). Treatment consisted of an arTMS course of 6 sessions/day (50 min inter-sessions intervals) over 5 days (on weekdays), thus totalling 30 sessions. After a 1-week gap, arTMS was followed by a tapering course of once-daily stimulation (minimum of 20, maximum of 25 sessions), 3-5 days per week, to decrease the odds of relapse.

*Baseline* assessments were completed during the week prior to arTMS initiation and consisted of a clinical assessment by trained research staff, including completion of the self-rated Beck Depression Inventory-II (BDI-II) and clinician-rated Hamilton Rating Scale for Depression 17-item (HRSD-17), cap fitting, and motor threshold calibration. Participants were reassessed at 3 subsequent timepoints: *post-acute* visits were conducted during the one-week gap following arTMS; *post-tapering* visits were conducted within 7 days of tapering termination; *1-month follow-up* visits were conducted 1-month (± 7 days) after the last tapering session. Participants who missed any sessions of the arTMS course were withdrawn. All participants were encouraged to complete the tapering course. Participants were asked not to change their medication regimen throughout the whole treatment period. The study timeline is illustrated in **Figure 3**.

**Figure 3:**
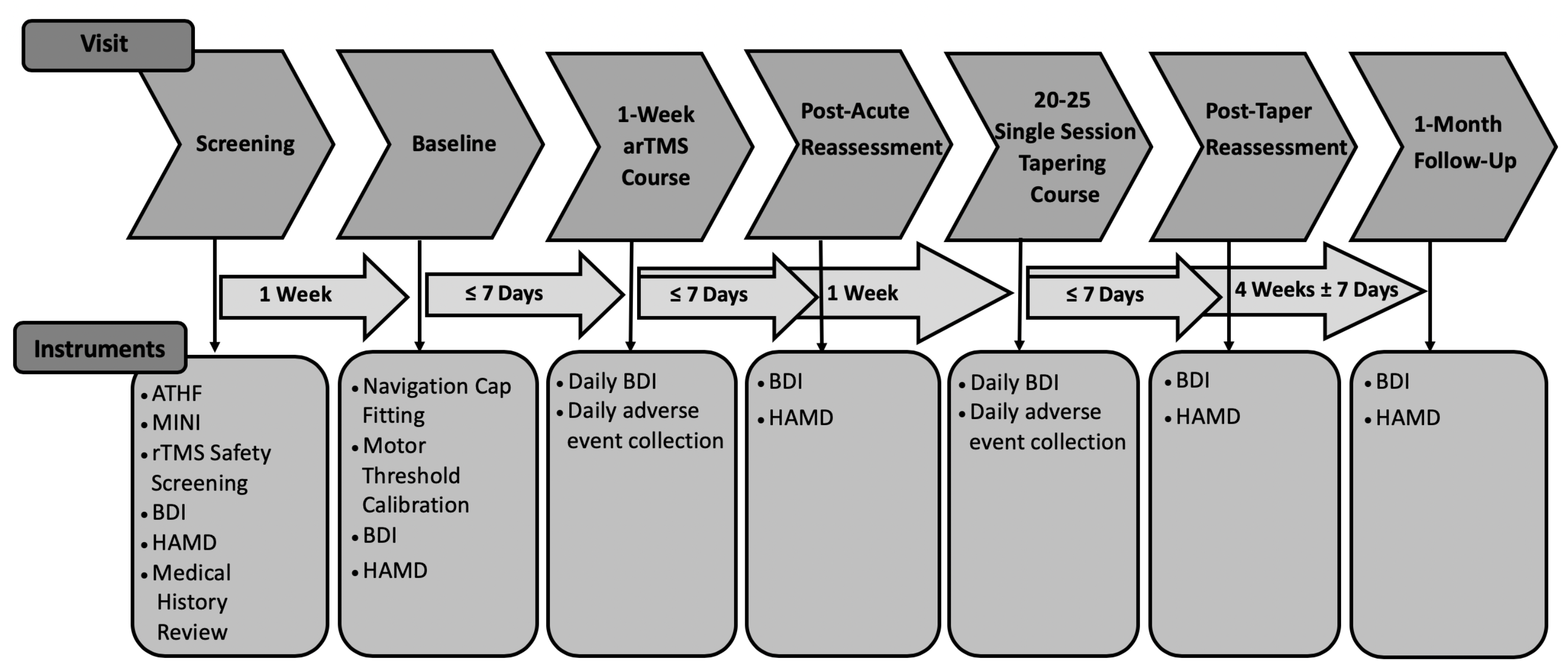
Study timeline - Treatment consisted of an arTMS course of 6 sessions/day (50 min inter-sessions intervals) over 5 days (on weekdays), thus totalling 30 sessions. After a 1-week gap, arTMS was followed by a tapering course of once-daily stimulation (minimum of 20, maximum of 25 sessions), 3-5 days per week, to decrease the odds of relapse. Baseline visits were conducted the week prior to arTMS initiation and consisted of a clinical assessment by a trained research staff, navigation cap fitting and motor threshold calibration. Patients were clinically reassessed at 3 subsequent timepoints: *post-acute* visits were conducted during the one-week gap following arTMS; *post-taper* visits were conducted within 7 days of tapering termination; *1-month follow-up* visits were conducted 1-month (± 7 days) after the last tapering session. arTMS = accelerated repetitive transcranial magnetic stimulation, ATHF = Average Antidepressant Treatment History Form, MINI = Mini International Neuropsychiatric Interview, BDI-II = Beck Depression Inventory-II, HRSD-17 = 17-item Hamilton Rating Scale for Depression

To study response trajectory during treatment days, participants also completed the BDI-II at the beginning of each treatment day before rTMS initiation, where they were queried about any adverse events. Self-rated pain intensity of the rTMS procedure was recorded on a verbal analog scale (VRS – from 0 [no pain] to 10 [intolerable pain]). Moreover, serious adverse events and reasons for treatment discontinuation were recorded when such events occurred. Stimulation intensity was adaptively titrated upward, aiming to reach the target intensity of 120% rMT on the first session of treatment, without exceeding maximum tolerable pain. We recorded the number of sessions required to reach 120% rMT.

### 2.3 Outcomes

Primary outcome measures were response and remission rates on the BDI-II. Secondary outcomes included score changes and percent improvement. These outcomes were also calculated on the HRSD-17. Response was defined as score reductions of ≥50% from baseline. Remission was defined as a score of ≤12 (Riedel et al., 2010) on the BDI-II and ≤7 on the HRSD-17 (Zimmerman et al., 2004). We also analyzed the outcome trajectories using the BDI-II.

### 2.4 Statistical analysis

Descriptive statistics were performed on baseline characteristics (age, sex, comorbid anxiety, age of onset of MDD, duration of current MDD episode, total lifetime number of antidepressant medication trials, total ATHF score and baseline BDI score) utilizing independent samples t-tests (two-tailed) for continuous variables, and Chi-square tests for categorical variables. We also performed repeated measures analyses of variance (ANOVA) on BDI-II score at different timepoints to assess the effect of the treatment through time. Planned repeated contrasts were used to make comparisons between the different evaluation times.

## 3. RESULTS

From March 18 to September 27, 2019, 57 participants with MDD were assessed for eligibility, 7 of whom were ineligible or declined to participate. 50 participants were thus enrolled and started the study. Of these, 2 discontinued treatment during arTMS because of lack of perceived efficacy and were lost to follow-up. 48 participants moved on to the tapering course and were thus included in the final analysis. Of these, 3 participants discontinued before having completed at least 20 sessions of the tapering course because of the lack of perceived efficacy. 45 participants thus completed the entire study, and 35 followed up at 1 month (**Figure 4**).

**Figure 4:**
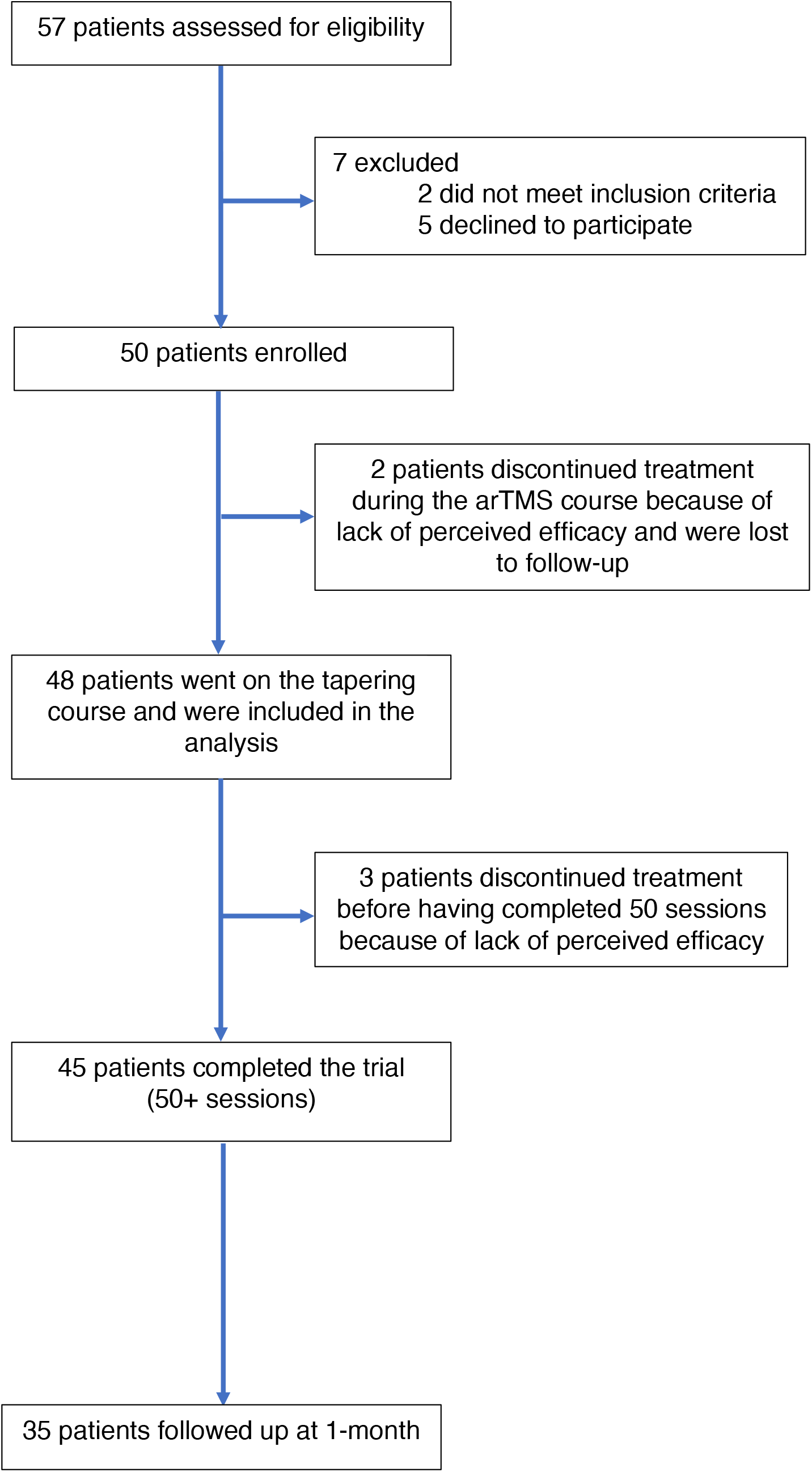
Trial CONSORT flow diagram - arTMS = accelerated repetitive transcranial magnetic stimulation.

**Table 1** provides the baseline characteristics of the study participants. Mean age was 41.8 ± 12.2, with 56.3% female participants. Mean age of depression onset was 23.8 ± 10.8 years old, with average length of current episode 51.7 ± 78.2 months. 70.8% of participants were receiving psychopharmacotherapy during the trial, with 58.3% taking at least one antidepressant during the study. Average Antidepressant Treatment History Form (ATHF) total score was 4.2 ± 4.3. Average number of trials on the ATHF in the current episode was 1.4 ± 2.6, with 39/48 (81.3%) of participants having had at least one adequate antidepressant trial in their current depressive episode. Comparing baseline characteristics variables between responders and non-responders did not yield any statistically significant differences (p ≥ .05).

**Table 1:**
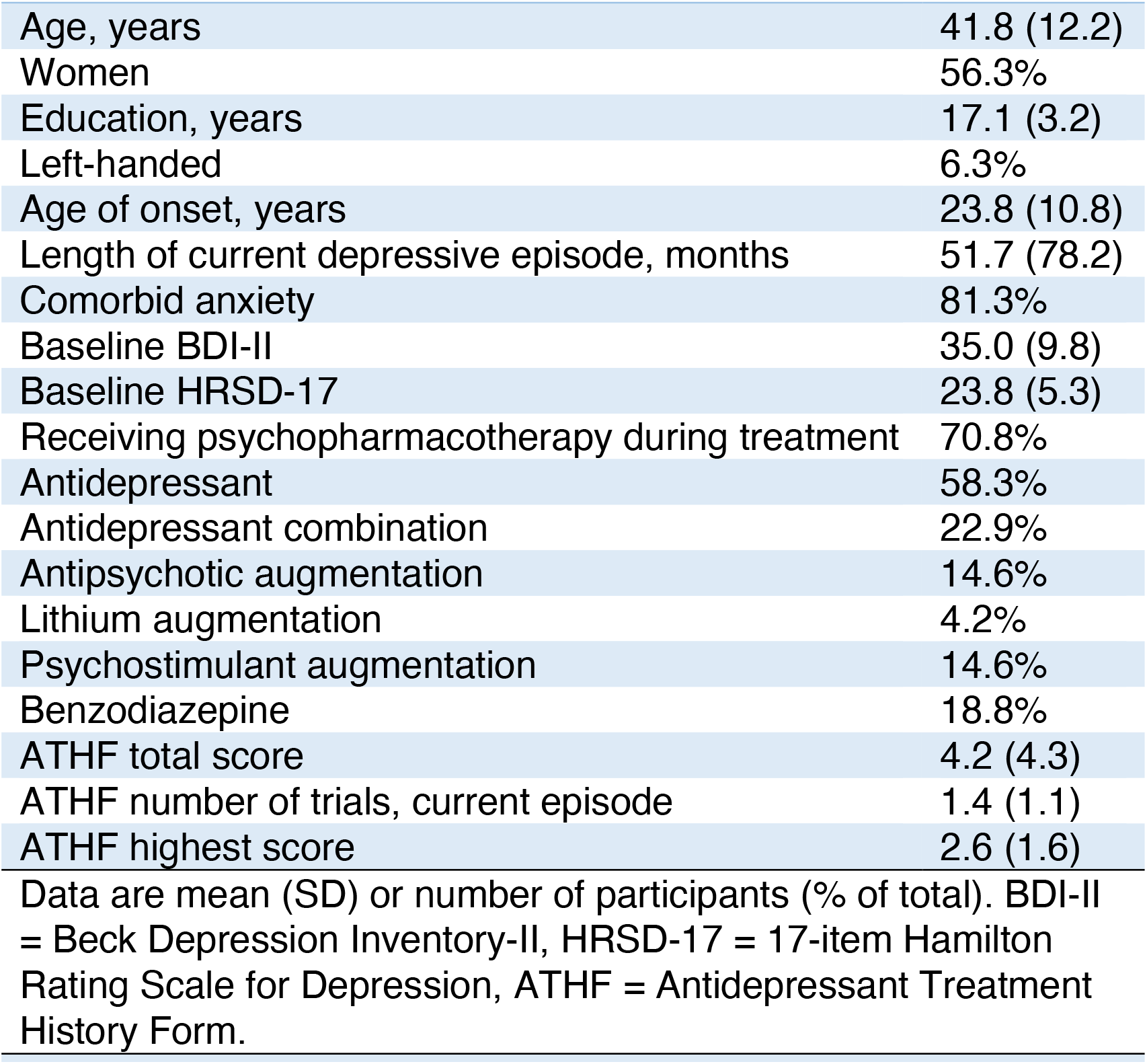
Baseline demographic and clinical characteristics (n = 48)

|  |  |
| --- | --- |
| Age, years | 41.8 (12.2) |
| Women | 56.3% |
| Education, years | 17.1 (3.2) |
| Left-handed | 6.3% |
| Age of onset, years | 23.8 (10.8) |
| Length of current depressive episode, months | 51.7 (78.2) |
| Comorbid anxiety | 81.3% |
| Baseline BDI-II | 35.0 (9.8) |
| Baseline HRSD-17 | 23.8 (5.3) |
| Receiving psychopharmacotherapy during treatment | 70.8% |
| Antidepressant | 58.3% |
| Antidepressant combination | 22.9% |
| Antipsychotic augmentation | 14.6% |
| Lithium augmentation | 4.2% |
| Psychostimulant augmentation | 14.6% |
| Benzodiazepine | 18.8% |
| ATHF total score | 4.2 (4.3) |
| ATHF number of trials, current episode | 1.4 (1.1) |
| ATHF highest score | 2.6 (1.6) |

Data are mean (SD) or number of participants (% of total). BDI-II = Beck Depression Inventory-II, HRSD-17 = 17-item Hamilton Rating Scale for Depression, ATHF = Antidepressant Treatment History Form.

Safety and tolerability outcomes are presented in **Table 2**. No serious adverse events (AE) were reported at any point of the study. One patient was withdrawn at session 51 since he reported visual symptoms suggestive of possible retinal detachment but was assessed and cleared by ophthalmology (subsequent diagnosis of migraine equivalent). Overall, 39.6% of participants reported at least one occurrence of an AE at some point during treatment, the most common one having been headache (22.9%). All AEs were reported exclusively during the arTMS course, with the exception of the previously mentioned participant with visual symptoms at session 51. Pain ratings went from 5.6 ± 2.0 on the first treatment, down to 1.9 ± 1.9 on the last treatment. The average rMT was 34.6 ± 5.7% of maximum stimulator output, resulting in a mean target stimulation intensity (120%) of 41.8 ± 6.8%. All participants were able to reach their target stimulation intensity, averaging 2.4 ± 3.0 sessions (1.2 ± 0.4 days) to do so, with 33/48 (68.8%) able to achieve it during their first session of treatment. Average total number of sessions was 54.1 ± 2.9, and mean overall treatment length, including the week gap between arTMS and tapering course, was 8.39 ± 0.93 weeks.

**Table 2:** Adverse events

|  |  |
| --- | --- |
| Serious AE | 0/48 (0.0%) |
| AE total | 19/48 (39.6%) |
| Headache | 11/48 (22.9%) |
| Fatigue | 7/48 (14.6%) |
| Nausea | 2/48 (4.2%) |
| Scalp tenderness | 3/48 (4.2%) |
| Hand twitching | 1/48 (2.1%) |
| Insomnia | 3/48 (6.3%) |
| Dizziness | 1/48 (2.1%) |
| Anxiety | 3/48 (6.3%) |
| Jaw pain | 1/48 (2.1%) |
| Visual symptoms | 1/48 (2.1%) |
| First treatment pain VRS | 5.6 (2.0) |
| Last treatment pain VRS | 1.9 (1.9) |
| Number of days to reach target stimulation intensity | 3.8 (6.3) |
| Number of participants (n=48) reporting adverse events (AE - %). For pain, data mean (SD). VRS = Verbal Rating Scale. |  |

**Table 3** presents the outcomes of interest. Regarding primary outcomes, response/remission rates on the BDI-II were 25.0%/16.7% *post-acutely*, 35.4%/27.1% *post-taper*, and 28.6%/22.9% at *1 month*. Regarding secondary outcomes, scores decreased from 35.0 (9.8 SD) at baseline down to 25.8 (12.2) *post-acutely*, to 22.2 (13.1) *post-tapering*, and 21.5 (12.1) at *1 month*. Percent improvement *post-acutely* was 27.4% (27.8% SD), 37.9% (33.0%) *post-tapering*, and 36.1% (34.7%) at *1 month*. On the HRSD-17, response/remission rates were 22.9%/6.3% *post-acutely*, 58.3%/37.5% *post-taper*, and 60.0%/37.1% at *1 month*. Scores decreased from 23.8 (5.3 SD) at baseline down to 16.3 (6.6) *post-acutely*, to 12.2 (8.0) *post-tapering*, and 11.3 (7.4) at *1 month*. Percent improvement *post-acutely* was 31.4% (23.8% SD), 48.6% (31.9%) *post-tapering*, and 52.4% (28.5%) at *1 month*.

Also, since we collected daily BDI-II during treatment days, we were able to assess trajectories of outcomes, presented in **Figure 5**. Overall, responders showed rapid improvement during the treatment week, having achieved response on average *post-acutely*, and continued to show slow but steady additional improvement up to the end of the tapering course. Repeated measures ANOVA showed significant changes of BDI-II scores over time (F 2.9;85.4 = 16.6, P < .001) with planned contrasts revealing a significant difference between baseline and day 2 (P = .007), day 4 and 5 (P = .029), day 5 and FU1 (P = .013), as well as between FU1 and FU2 (P = .002) and no significant change between other assessments. A similar pattern was observed for the HDRS-17 (F 2.2;74.1 = 68.4, P < .0001). There was no significant difference between response and remission rates on the BDI-II and HRSD-17 throughout all measurement times (FU1, FU2, FU3).

**Table 3:** Outcomes of interest

|  |  |
| --- | --- |
| BDI-II |  |
| Response *post-acute* | 12/48 (25.0%) |
| Response *post-taper* | 17/48 (35.4%) |
| Response *1-month* | 10/35 (28.6%) |
| Remission *post-acute* | 8/48 (16.7%) |
| Remission *post-taper* | 13/48 (27.1%) |
| Remission *1-month* | 8/35 (22.9%) |
| Score baseline | 35.0 (9.8) |
| Score change *post-acute* | 25.8 (12.2) |
| Score change *post-taper* | 22.2 (13.1) |
| Score change *1-month* | 21.5 (12.1) |
| Percent improvement *post-acute* | 27.4% (27.8%) |
| Percent improvement *post-taper* | 37.9% (33.0%) |
| Percent improvement *1-month* | 36.1% (34.7%) |
| HRSD-17 |  |
| Response *post-acute* | 11/48 (22.9%) |
| Response *post-taper* | 28/48 (58.3%) |
| Response *1-month* | 21/35 (60.0%) |
| Remission *post-acute* | 3/48 (6.3%) |
| Remission *post-taper* | 18/48 (37.5%) |
| Remission *1-month* | 13/35 (37.1%) |
| Score baseline | 23.8 (5.3) |
| Score change *post-acute* | 16.3 (6.6) |
| Score change *post-taper* | 12.2 (8.0) |
| Score change *1-month* | 11.3 (7.4) |
| Percent improvement *post-acute* | 31.4% (23.8%) |
| Percent improvement *post-taper* | 48.6% (31.9%) |
| Percent improvement *1-month* | 52.4% (28.5%) |
| Data are mean (SD). For remission and response rates, data are n (% of participants assessed). BDI-II = Beck Depression Inventory-II, HRSD-17 = 17-item Hamilton Rating Scale for Depression. |  |

**Fig. 5.**
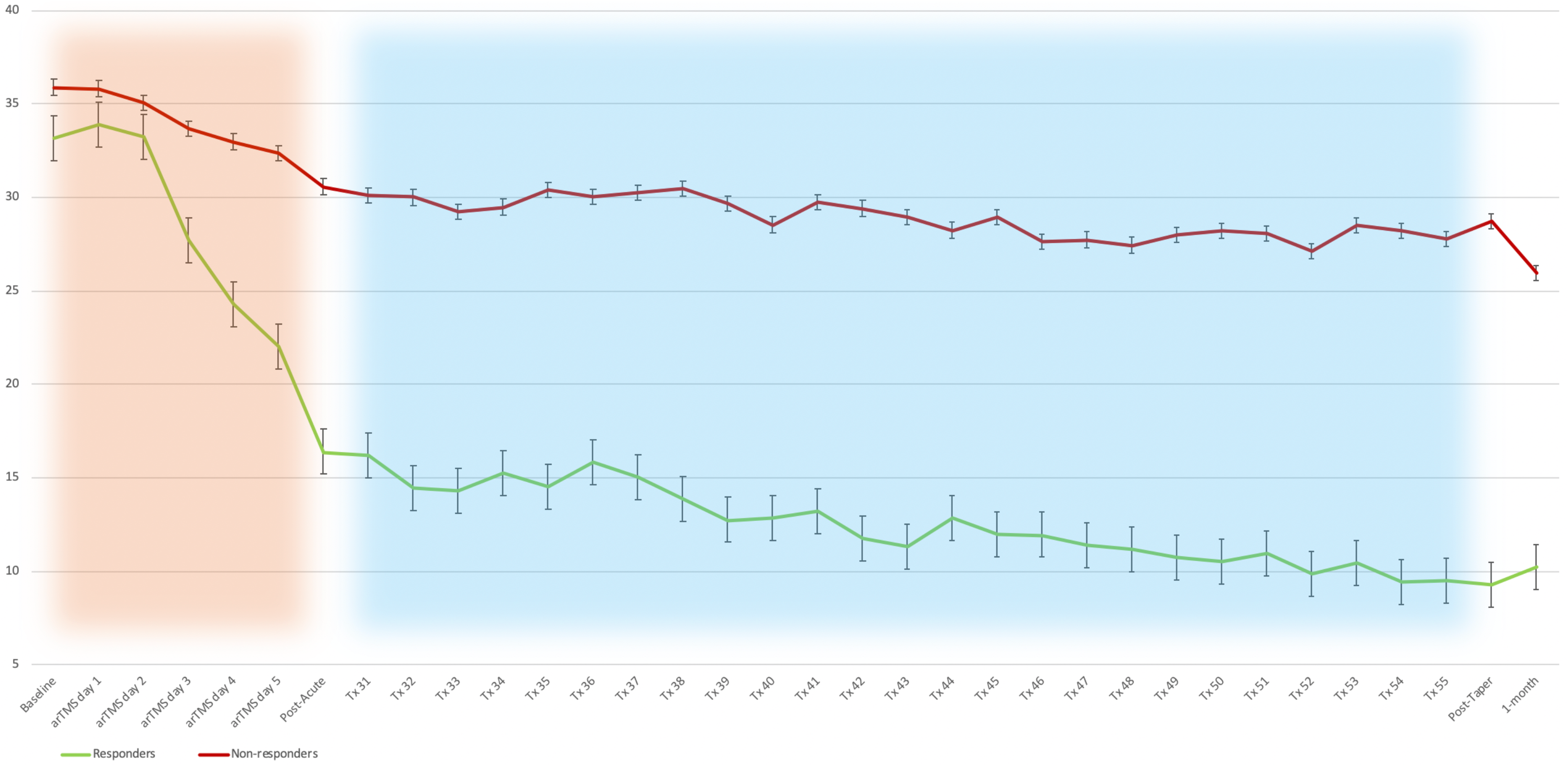
Trajectories of improvement on the BDI-II. Responders showed rapid improvement during the accelerated course, having achieved response on average response post-acutely, and continued to show slow but steady additional improvement up to the end of the tapering course. Use of background shading delineates arTMS from tapering course. BDI-II = the Beck Depression Inventory – II, arTMS = accelerated repetitive transcranial magnetic stimulation, Tx = treatment (rTMS session)

## 4. DISCUSSION

We present preliminary data for a novel rTMS technique optimized for widespread use. Currently, most guidelines recommend rTMS use solely in treatment-resistant depression (TRD). Specifically, the Canadian Network for Mood and Anxiety Treatments (CANMAT) recommends use in MDD participants who have failed to respond to at least 1 antidepressant (Milev et al., 2016), a decision mirrored in other international guidelines (Lefaucheur et al., 2020; Perera et al., 2016). However, access could potentially be broadened if treatments could be delivered safely in more centres, at higher volumes and lower costs, with a faster onset of response. Our study thus attempted to resolve these issues by offering several technical refinements.

First, treatment was delivered in an open-room setting, allowing one or two technicians to treat up to 4 participants in parallel, compared to the usual one-on-one approach. Even though such a setting could bring about some concerns about technician fatigue and negative patient feedback, this was not reported. We also did not observe any negative interactions between participants that could have jeopardized treatment outcome, and several participants anecdotally gave positive feedback about the more flexible scheduling of treatment sessions enabled by the open setting. Overall, this approach merits qualitative assessment with standardized feedback questionnaires for both technicians and participants. If successful on a larger scale, we believe that this approach may reduce staffing demands, increase clinic capacity and flexibility, and improve cost and accessibility. Given the current COVID-19 pandemic, reduce staffing might be a potential advantage in order to observe physical distancing measures, but it would need to be conjugated with stringent infection control procedures.

One innovation that permitted such a treatment setting was the design and use of our novel AAAs. Standard articulated arms can be more expensive, while offering a limited range of motion. Our AAAs are made of widely available camera tripod equipment and have been both functional and reliable, with no material malfunctions observed. Their durable yet bendable extended branch allowed for the treatment technician to easily and accurately position the coil without tedious joint adjustments. Their anchoring to the treatment chair also eliminated any standalone equipment or cart, creating a more spacious, barrier-free clinic. The mechanical advantage of the anchoring point gives the patient the flexibility to raise or lower the incline of their treatment chair without placement adjustment, as the coil is now anchored to the shifting point of reference, thereby maintaining the coils’ targeted area of focus. This custom solution decreased operational complexity, allowing for better patient care, ease of use for the technician, and more efficient use of space and equipment. Similar coil positioning equipment would be relatively straightforward for rTMS manufacturers to adopt in future.

The use of 1 Hz stimulation is another major component allowing for simplified rTMS delivery. Firstly, 1 Hz rTMS is the medically safest stimulation pattern available, having even shown anti-seizure properties in epileptic patients (Sun et al., 2012). Further, there exists empirical evidence that 1 Hz rTMS is better tolerated than HF protocols (Kaur et al., 2019), and also shows similar efficacy (Brunoni et al., 2017; Cao et al., 2018; Fitzgerald et al., 2019). With preliminary data potentially showing a higher safety and tolerability profile, LF 1 Hz could be considered in the future as a first line rTMS protocol. Beyond these factors, the greatest advantage of 1 Hz rTMS lies in the low cost of the equipment it requires. HF rTMS require higher-cost stimulators, with expensive cooling systems. 1 Hz rTMS on the other hand can be delivered on more simple stimulators with coils that do not require cooling. This setup is more affordable than the ones required for HF, with the possibility of additional cost reductions in the future through higher volume use.

Parabolic coils also allow simplification of rTMS delivery. We previously published a case report on their use (Miron et al., 2019), delineating their potential advantage over Fo8 coils. Indeed, large parabolic coils may require less precise placement given their large electromagnetic field compared to the target brain region – a factor also present with larger helmet-shaped coils (Levkovitz et al., 2015). Also, their central opening allows direct visualization of underlying landmarks, resulting in easier accurate placement over the marked target. We also hypothesized that, given their possibly weaker stimulation of the DLPFC owed to the aforementioned opening, parabolic coils may function by stimulating adjacent neural substrates, such as the orbitofrontal cortex (OFC). Our group also previously published a case series on 1 Hz OFC, which is theorized to be involved in depression (Downar, 2019; Rao et al., 2018; Rolls, 2016).

Another novel aspect of our study was the piloting of accelerated 1 Hz rTMS. We are aware of only one small (n = 7) negative trial with limited stimulation course and duration (18 sessions over 10 days) (Tor et al., 2016). Our results are encouraging, but overall limited response and remission rates after the arTMS course would suggest that increasing the number of sessions per day and the number of pulses delivered at each session may be necessary (Baeken, 2018). Still, analyses showed a rapid effect during the accelerated phase, which supports the idea that intensive protocols with a high number of daily sessions could be an important ingredient to increase response. Indeed, a recent arTMS pilot (n = 6) with highly refractory MDD patients using intermittent theta-burst stimulation (iTBS), 1800 pulses/session, 10 sessions per day over 5 days, reported remission rates of 66% (Williams et al., 2018). A larger follow-up study (n = 21) reported remission rates of up to 90% (Cole et al., 2020). The rapid improvement in responders *post-acutely* (**Figure 5**) is encouraging, showing that it is indeed possible to rapidly accelerate response to 1 Hz stimulation in some individuals. This could justify further the study of this rTMS technique in more severe inpatient cases, where suicidality is of concern and rapid response is at a premium. Overall, final outcomes *post-taper* compared favorably to another large trial (Blumberger et al., 2018), as well as a landmark meta-analysis (M T Berlim et al., 2012), which may be explained by the overall high number of sessions. Supporting this observation, a recent study that offered up to 21 weeks of rTMS reported remission rates of 72% (Stubbeman et al., 2018). Since there is a lack of data about 1 Hz arTMS alone without a subsequent tapering period, we are unable to determine the impact or the necessity of such an extension. It may well be possible that 1 Hz arTMS has a delayed effect, where participants would have kept on improving without the extension course, as suggested in other arTMS protocols (Sonmez et al., 2019). Conversely, high relapse rates following arTMS are also of concern (Caulfield, 2019; Williams et al., 2018).

This preliminary study has several limitations. Of primary concern is the open-label design and the absence of a sham control arm. It is to be expected that the various estimates of effectiveness might be higher than what would be obtained in a randomized, sham-controlled trial, which would represent the next logical step. In addition, the naturalistic approach of the study does not allow estimates of efficacy, but only effectiveness. Also, we did not qualitatively assess our open-room setting, as discussed previously. As a further matter, the weaker central field in the parabolic coils used may have under-stimulated the DLPFC (Miron et al., 2019); this may be correctable in future by moving the central opening to an intermediate mark between F4, Fz and Cz. Due to their less widespread use in clinical settings, results with 1 Hz rTMS on parabolic coils may not generalize to more commonly used Fo8 coils. The 1 Hz protocol used, although highly tolerable for participants, may also have been less effective due to its limited amount of pulses per session. Given that increased number of pulses has been associated with increased response in 1 Hz rTMS in some studies (Marcelo T Berlim et al., 2012), it may be advantageous to increase the number of pulses within the same time frame (e.g., 600 pulses in 10 min). Finally, although we did not require participants to meet the usual requirement of TRD in our trial, the majority (>80%) of participants had failed at least one adequate trial. We also did not have a minimal cut-off regarding depression severity on the mood scales for study inclusion, but average baseline scores on the BDI-II was in the severe range. Average HRSD-17 score was also higher than in a recent large trial from our group (Blumberger et al., 2018).

Given its established efficacy in MDD and lower side-effect profile compared to medication, efforts should be made to decrease costs and complexity associated with rTMS, which could increase accessibility to the patient population. If decreased complexity can bring down costs to a level comparable to some medication regimens, the kind of techniques demonstrated here may be suitable for safe use in a wider range of settings, including more community-based approaches. If the safety, tolerability and efficacy of such protocols can be confirmed in randomized controlled trials, it is possible to imagine rTMS becoming eventually a viable first-line treatment option for MDD.

## Data Availability

Upon reasonable request

## CONTRIBUTIONS

**JPM** designed the study, was responsible for data collection, analyzed the data, and wrote the manuscript. **HV, LF, MH** participated in the study design, data collection, and reviewed the manuscript. **FM** and **VDJ** participated in data analysis and reviewed the manuscript. participated in data collection and reviewed the manuscript. **SD, RZ, JS, AM, DMB, ZJD** and **FVR** participated in the study design and reviewed the manuscript. **JD** provided resources, supervised all phases of the study and reviewed the manuscript.

## ACKNOWLEDGEMENT

**JPM** would like to thank the Brain & Behavior Research Foundation and the Branch Out Neurological Foundation for their financial support of this project. We would like to thank Terri Cairo, Julian Kwok, Meaghan Todd, Nuno Ferreira, Thomas Russell and Eileen Lam for their involvement and organizational support throughout this project. This manuscript has been released as a pre-print at medRxiv (Miron et al., 2020).

